# The socio-economic determinants of COVID-19: A spatial analysis of German county level data

**DOI:** 10.1101/2020.06.25.20140459

**Authors:** Andree Ehlert

## Abstract

The study explores the influence of socio-economic variables on case and death rates of the COVID-19 pandemic in Germany until mid-June 2020. It covers Germany’s 401 counties by multivariate spatial models that can take into account regional interrelationships and possible spillover effects. The case and death rates are, for example, significantly positively associated with early cases from the beginning of the epidemic, the average age, the population density and the number of people employed in elderly care. By contrast, they are significantly negatively associated with the density of schoolchildren and infant care as well as the density of doctors. In addition, for certain variables significant spillover effects on the case numbers of neighbouring regions could be identified, which have a different sign than the overall effects and thus give cause for further analyses of the mechanisms of action of COVID-19 infections. The results complement the knowledge about COVID-19 infection beyond the clinical risk factors discussed so far by a socio-economic perspective. The findings can contribute to the targeted derivation of political measures and their review, as is currently being discussed in particular for the tourism and education sectors.

## 1 Introduction

Since its outbreak in December 2019 in Wuhan Province, China, the respiratory disease COVID-19 has developed in just a few weeks into a pandemic with currently (22 June 2020) about 9 million infections worldwide and about 469,000 deaths. Many countries have responded with a comprehensive lockdown (including curfews and school closures), in particular to avoid overloading the health system. Although the initially exponential growth in case numbers has now slowed down in many countries, there is still no vaccination and so far no herd immunity. Therefore, the concrete danger of further waves of infection exists.

In Germany the first confirmed cases were reported at the end of January 2020. The number of new infections per day rose to more than 100 at the beginning of March 2020 and reached a maximum of about 7,000 at the end of March. The initial cases were mainly attributed to imported clusters spread at major events (e.g. sports, carnival).

The social and economic consequences of the lockdown measures taken in almost all countries to contain the pandemic are serious and their long-term development is not yet foreseeable. This increases the pressure for empirical evaluation of previous contact restrictions and, hence, for the development and implementation of targeted and efficient future political measures. While clinical and epidemiological research is currently discussing person-specific risk factors for infection or probability of survival, which often have a strong ad hoc influence on government lockdown measures, there are so far few results on the question of which socio-economic and region-specific factors (after controlling the initial case numbers of a region) are associated with the spread of COVID-19. This approach is about the manifold possibilities to fight the epidemic with measures that lie outside the health sector in the narrow sense [6].

Knowledge of such socio-economic risk factors would have far-reaching social relevance, since target-group-specific adaptation of contact restrictions can reduce the social and economic costs of the measures. In particular, the probability of infection and survival may also be directly affected, if, for example, knowledge of the factors associated with an infection or survival allows the identification of subgroups or regions that are particularly worthy of protection. Conversely, of course, this also means a limitation of the present analysis. No direct, individual risk factors (such as those currently motivated to protect certain population groups via previous illnesses, the job or the family situation) can be identified. Clearly, it is not possible to map the behaviour of individuals in an ecological study (such as their contacts). Rather, an ecological analysis is more concerned with the causes of the causes which favour the spread of COVID-19 through socio-economic factors at regional level in order to gain possible influence on the actual mechanism of action. With regard to the mechanisms of action of the infection, it can be assumed that persons of low socioeconomic status are simply more exposed to the clinical infectious factors of COVID-19. However, this discussion is outside the scope of the paper, see [28, 16] for details.

The hypotheses concerning socio-economic factors influencing COVID-19 are manifold. For example, the average age of the population, regional vaccination rates against tuberculosis, climatic factors, or the way schools and childcare facilities are managed, have recently been discussed in the daily press in terms of their impact on the spread of COVID-19. With this ecological analysis, further socio-economic factors at the level of the 401 administrative districts (counties) in Germany (whose infection incidence currently varies considerably) will be analysed with the help of spatial statistical methods. Besides the explorative character of this analysis (at the current state of research), three questions in particular are to be answered empirically: First, what possible influence do schoolchildren and children in day care have on the occurrence of infection? Second, can the thesis of the spread of COVID-19 via institutions for care of the elderly be empirically confirmed at regional level? Third, what influence do spillover effects, i.e. the interlinked spread of COVID-19 between neighbouring regions, have on the infection rate? Other factors which have not yet been quantitatively investigated and which can be answered with the data of this study include, for example, the question of the influence of the size of the dwelling (regarding the question of frequency of contacts), tourism (supra-regional spread) or the sectoral structure (contacts in the office or on the building site).

To answer these questions, the case numbers and death rates at the district level provided for Germany by the Robert Koch-Institut (RKI) are associated with socio-economic indicators at the district level, such as education, income, unemployment, information on health status and health care, school, infant care, etc. Spatial econometric models such as the spatial autoregressive model (SAR) or the spatial error model (SEM) are used for our analysis, which take into account the distance between districts using a weight matrix. These models can reduce possible bias of an ordinary least squares (OLS) estimation and/or increase the efficiency of the estimation.

This paper complements the existing literature in at least three areas. Firstly, the literature on COVID-19 tends to focus on clinical studies to identify individual risk factors and develop curative treatments, see e.g. [33]. However, the contribution of this study lies precisely in the identification of key socio-economic factors and the possible regional mechanisms of action through which COVID-19 spreads. Knowledge of these factors and the ways in which they are spread is crucial, since at the current state of research it seems not to be possible to contain the pandemic by medical means alone. Examples from the literature on the analysis of socio-economic risk factors of health status are numerous. As a classic example, [10] deals with the various, also socio-economic, determinants of individual health. For infectious diseases such as pneumonia and influenza, [11] has investigated the influence of socio-economic factors in addition to individual risk. socio-economic risk factors for HIV were analysed by [1], for pertussis by [14], for salmonella infections by [31] and for H1N1 pandemic mortality in the USA by [24], to name only some examples.

Secondly, socio-economic risk factors in relation to COVID-19 have so far been analysed rather for regions outside Germany or in an international comparison of countries. Examples are [21] for 20 European countries, which points to a positive influence of high social activity and high population density on COVID-19 infections, or [28, 29] for a worldwide analysis of risk factors based on country data. Examples of regional analyses in Spain are [22], for an analysis at municipal level within Catalonia [20], for France [12], for Iran [26], for China [25] or [4] for Italy, to name a few. For the analysis at the district level within Germany, [23] finds significantly negative effects especially on income and education (for a later phase of the pandemic in May 2020). In a spatial regression discontinuity analysis [3] investigate the influence of the tuberculosis vaccination along the former inner-German border under consideration of numerous socio-economic factors at the district level. A time series approach taking into account regional factors such as age and population density was carried out by [19] and aims to explain the decline in infection rates in the later phase of the epidemic.

Thirdly, however, in the above analyses (with the exception of [5]) no spatial economtric models are applied. The latter take into account the distance of the districts from each other and thus the influence of possible spatial spillover effects. Outside Germany, similar approaches have been applied by [27] for spatial models (without spillover effects) at the European country level. For 31 regions in China [13] analyses the spatial spillover effects for the COVID-19 case numbers, but without considering socio-economic covariates.

The rest of the paper is structured as follows. Section 2 details the data and methods used. Results are presented in Section 3. Section 4 provides a discussion and concludes.

## 2 Methods

### 2.1 The data

Our analysis is a retrospective ecological study on the level of the 401 administrative districts and district-free cities (counties) in Germany, whose population ranges from about 34,000 (Zweibrücken) to about 3,664,000 (Berlin). It should be emphasized again that this is not a clinical study on the level of single individuals, so that only statements on the population level can be made. However, this study design is well established in the health sciences and provides very practice-oriented results, especially for questions in the areas of health services research, public health and epidemiology. See also [18] for a detailed discussion.

As outcome variables the case numbers and deaths at the district level, which are currently updated daily by the RKI, are used. The cut-off date for this analysis is 15. June 2020. The data are based on reports from the local health authorities to the RKI, which is why there may be a certain delay in reporting (especially on weekends). As officially recorded COVID-19 cases, the figures are a lower limit of the actual number of cases present (dark figure).

The socio-economic covariables at the district level come from two public sources, namely the Federal Institute for Research on Building, Urban Affairs and Spatial Development (BBSR) and the Central Research Institute of Ambulatory Health Care in Germany (ZI). Table 1 provides an overview of the data.

**Table 1:**
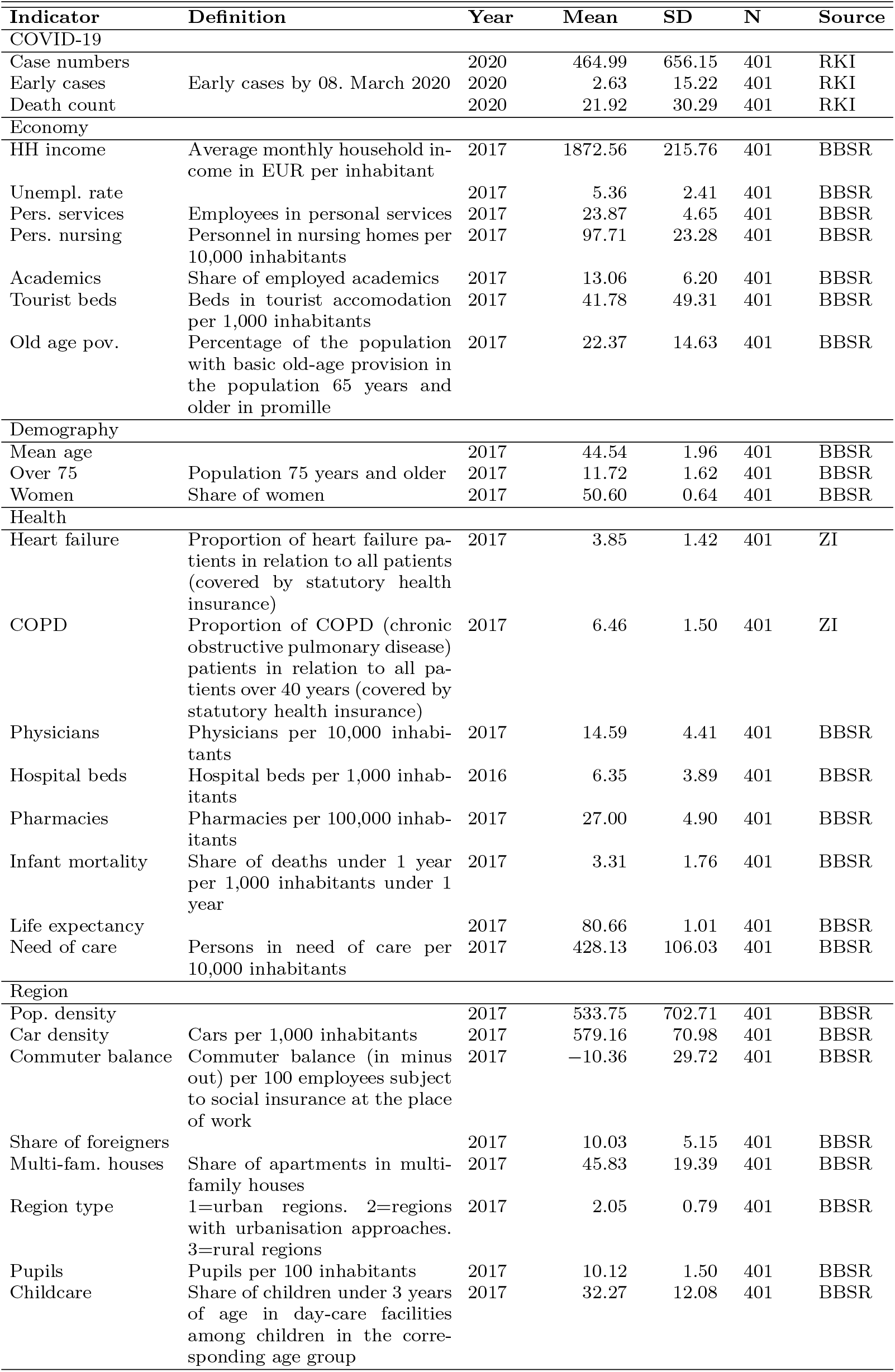
Basic sample characteristics for the outcome and main explanatory variables at the county level.

In selecting the socio-economic variables in Table 1, we followed existing ecological studies on respiratory diseases. For example, the influence of income and education variables as well as the region type on COVID-19 infections is discussed in [23], whereas [17] also cover the employment level. Demographic variables, population density and information on health status are considered in [3]. The physician density is brought into play in [25] in connection with COVID-19 infections in China. A very similar selection and grouping of socio-economic factors (but at the level of nation states) on COVID-19 infections is used in [28].

The actual clinical mechanisms of action behind the socio-economic factors are complex. For example, they can act simultaneously at the individual and population level and their direction is often unclear a priori. In the case of the economic status of a region, for example, the better infrastructure available to bridge a lockdown or further contact restrictions can be seen as an effect that slows down the spread of infection. The same applies to better medical treatment. On the other hand, a region’s higher economic output generally increases the networking and thus the frequency of contact between individuals, which can have an increasing effect on the number of infections. Also, with higher economic performance, more resources are available for testing for COVID-19, so that this alone can also result in a positive association.

### 2.2 Econometric methods

In contrast to a standard linear model, in our analysis the spatial distance of the observation units (counties) from each other will be taken into account. This can lead to spillover effects of the dependent variable in neighbouring regions or to a spatial autocorrelation of the residuals. To reflect this, different models are proposed in spatial statistics. Common to all of them is the description of the neighbourhood relations via a so-called spatial weighting matrix (symmetrical *N × N* matrix). For this purpose, the geocodes (longitude and latitude of the centre of the districts) provided by the provider Opendatasoft (under the creative commons licence) were used.^1^ The Stata command spmat [7] creates an inverse distance matrix from the coordinates, so that according to Tobler’s law [30] regions closer to each other receive a higher weight.

There are then essentially two approaches to spatial autocorrelation in the dependent variable or error terms, namely SAR and SEM, as well as numerous combinations and variations thereof. The technical details shall be omitted here, with reference to the excellent presentation in [9] or especially for an application in the context of COVID-19 in [13]. It is important, however, to note that in the presence of the SAR model the estimates of an OLS model may be biased and that the true effect of an independent variable is reflected by direct and indirect spillover effects (and, together, total effects). We will come back to this when discussing the results in Section 3. In a SEM model, there are no spatial spillover effects, but due to the spatial structure in the error terms, simple OLS estimation might be inefficient. We will consider a third model in our analysis, the so-called SAC as a combination of spatial autocorrelation in the dependent variables and in the errors, and refer to [9] for the technical details here as well.

## 3 Results

### 3.1 Case numbers

The econometric analysis was performed with the Stata commands spreg [8] and xsmle [2]. As a first step, it should be checked whether significant spatial dependency exists at all compared to an OLS model and which of the three proposed models best describes the data. Therefore, as suggested in [9], a likelihood ratio (LR) test of the models against an OLS model is performed. The results are given in the penultimate line of Table 2. Apparently the OLS model can be rejected against all three spatial models at least to the 5% level. Of the latter, both SAR (1% level) and SEM (5% level) can be rejected against the SAC (which are nested models by design).

**Table 2:** COVID-19 case numbers: Coefficient estimates for the SAR, SEM and SAC models discussed in Section 2.2. The significance level symbols are † for 10%, ∗ for 5%, and ∗∗ for 1 %

| Variable | SAR |  | SEM |  | SAC |  |
| --- | --- | --- | --- | --- | --- | --- |
|  | Coeff. | (Std. Err.) | Coeff. | (Std. Err.) | Coeff. | (Std. Err.) |
| Early cases | 7.04** | (1.25) | 6.82** | (1.24) | 6.66** | (1.22) |
| Economy |  |  |  |  |  |  |
| HH income | 0.24 | (0.17) | 0.15 | (0.17) | 0.18 | (0.17) |
| Unempl. rate | -12.34 | (22.69) | -19.15 | (22.55) | -15.55 | (28.70) |
| Pers. Services | -4.44 | (7.13) | -8.43 | (7.29) | -5.72 | (8.39) |
| Pers. nursing | 3.26* | (1.42) | 3.17* | (1.40) | 3.20* | (1.32) |
| Academics | 26.89** | (8.64) | 25.74** | (8.61) | 24.55† | (13.56) |
| Tourist beds | -0.65 | (0.55) | -0.59 | (0.55) | -0.61 | (0.66) |
| Old age pov. | 5.75 | (3.69) | 5.80 | (3.66) | 5.46 | (3.45) |
| Demography |  |  |  |  |  |  |
| Mean age | 130.33* | (51.21) | 134.12* | (52.33) | 159.12 | (143.59) |
| Over 75 | -141.15** | (51.21) | -149.03** | (53.11) | -165.25 | (123.51) |
| Women | 50.68 | (44.97) | 70.42 | (45.62) | 78.67 | (.) |
| Health |  |  |  |  |  |  |
| Heart failure | 10.79 | (24.67) | 5.68 | (24.72) | -6.28 | (29.84) |
| COPD | -26.29 | (20.81) | -17.50 | (21.64) | -14.06 | (22.25) |
| Physicians | -31.71* | (12.38) | -37.08** | (12.35) | -39.08** | (14.83) |
| Hospital beds | -6.27 | (9.15) | -2.85 | (8.95) | -5.46 | (8.60) |
| Pharmacies | 2.44 | (6.25) | 6.00 | (6.40) | 1.97 | (6.39) |
| Infant mortality | -0.73 | (11.07) | -0.53 | (10.85) | 0.13 | (11.08) |
| Life expectancy | 153.93** | (40.36) | 130.35** | (41.94) | 147.75** | (56.97) |
| Need of care | 0.41 | (0.42) | 0.35 | (0.43) | 0.43 | (0.48) |
| Region |  |  |  |  |  |  |
| Pop. density | 0.34** | (0.07) | 0.37** | (0.07) | 0.39** | (0.07) |
| Car density | -0.37 | (0.55) | -0.27 | (0.54) | -0.38 | (0.53) |
| Commuter balance | 1.34 | (1.45) | 0.96 | (1.45) | 1.42 | (1.48) |
| Share of foreigners | 6.21 | (10.23) | 0.94 | (10.24) | 4.39 | (10.31) |
| Multi-fam. houses | -3.11 | (3.65) | -1.68 | (3.69) | -1.32 | (4.37) |
| Region type | 11.43 | (39.20) | -1.09 | (40.68) | 8.27 | (40.29) |
| Pupils | -39.85* | (19.37) | -41.77* | (18.50) | -35.34† | (20.85) |
| Childcare | -7.84† | (4.67) | -5.68 | (4.84) | -8.46 | (7.58) |
| Constant | -18 035.32** | (4209.79) | -17 033.79** | (4529.10) | -19 835.75 | (.) |
| lambda | -0.46** | (0.15) |  |  | -0.50** | (0.18) |
| rho | 0.45** | (0.01) | 0.84** | (0.24) |  |  |
| sigma2 | 124 560.24** | (8817.05) | 122 414.75** | (8656.93) | 119 483.46** | (8817.90) |
| Log likelihood | -2922.85 |  | -2921.59 |  | -2918.86 |  |
| LR chi2(2/3) (OLS) | 8.71* |  | 11.22** |  | 16.68** |  |
| LR chi2(1) (SAC) | 7.97** |  | 5.46* |  |  |  |

The estimated coefficients in Tab. 2 show a rather uniform picture across the three spatial models with regard to their significance and sign. The individual independent variables are grouped in the left column as in Table 1, the estimated values with the corresponding standard errors in brackets are shown in the following columns for the respective models. Dummies for the 16 federal states were included in the model (estimates not shown) to check for the potential influence of country-specific lockdown measures and their different time frames.^2^ In detail, it can be seen that, as expected, the early case numbers have a significant (1% level) positive influence on case numbers (as of 15. June 2020). With regard to the economic variables, in contrast to what is expected (according to the media discussion), there is no significant negative influence of the income variable and no significant positive influence of the unemployment rate. Here, it is also important to point out once again that the results of an ecological study do not necessarily have to correspond with the infection incidence at the individual level, since it is a snapshot of population-related processes that are recorded here. However, similar results were reported in [28, 20], for example, and (in contrast to the prevalence of long-term widespread diseases) were attributed to the fact that the spread of the virus is favoured by high economic activity. However, it must be mentioned that the (not significant) negative coefficient of persons in personal service occupations would rather contradict this hypothesis.

Interesting with regard to our second research question in Section 1 is especially the positive influence of employees in the nursing professions across all three models (5% level). Apparently, even after controlling for the number of people in need of care (see further down in the table, not significant), the infection rate seems to be influenced by the number of people employed in care, which would confirm the current media discussion empirically consistent. On the other hand, the tourist bed sector cannot be significantly associated with the infection rate in our models, although numerous channels of action are conceivable even after the lockdown measures have come into effect. It is possible, however, that these have already manifested themselves in the early case numbers and are therefore largely covered by this variable.

The demographic variables reflect the fact that a rising average age has a significantly positive (5% level, except for SAC) influence on the number of cases, whereas the proportion of persons over 75 years seems to have a significantly negative (1% level, except for SAC) influence. This seems to contradict the media presentation that especially elderly people are affected by COVID-19 infections (which seems to apply even to the death figures with regard to Table 3, see below). However, this result can be empirically confirmed for example by [20] for Catalonia. As an explanation, it can again be cited that a high proportion of older persons tends to indicate a regionally lower level of economic and social activity, thus limiting the spread of the virus.

**Table 3:** COVID-19 death count: Coefficient estimates for the SAR, SEM and SAC models discussed in Section 2.2. The significance level symbols are as before.

| Variable | SAR |  | SEM |  | SAC |  |
| --- | --- | --- | --- | --- | --- | --- |
|  | Coeff. | (Std. Err.) | Coeff. | (Std. Err.) | Coeff. | (Std. Err.) |
| Early cases | 0.29** | (0.07) | 0.27** | (0.07) | 0.27** | (0.07) |
| Economy |  |  |  |  |  |  |
| HH income | 0.00 | (0.01) | 0.00 | (0.01) | 0.00 | (0.01) |
| Unempl. rate | -0.22 | (1.36) | -0.52 | (1.35) | -0.46 | (1.59) |
| Pers. Services | -0.35 | (0.43) | -0.54 | (0.44) | -0.42 | (0.43) |
| Pers. nursing | 0.27** | (0.08) | 0.29** | (0.08) | 0.29** | (0.09) |
| Academics | 1.05* | (0.52) | 1.05* | (0.51) | 0.96† | (0.54) |
| Tourist beds | -0.03 | (0.03) | -0.03 | (0.03) | -0.03 | (0.03) |
| Old age pov. | 0.34 | (0.22) | 0.36† | (0.22) | 0.35 | (0.22) |
| Demography |  |  |  |  |  |  |
| Mean age | 5.98† | (3.07) | 5.61† | (3.03) | 6.32* | (2.80) |
| Over 75 | -7.01* | (3.07) | -6.82* | (3.05) | -7.08** | (2.63) |
| Women | 3.93 | (2.69) | 4.76† | (2.68) | 4.88 | (5.37) |
| Health |  |  |  |  |  |  |
| Heart failure | 0.39 | (1.48) | 0.03 | (1.48) | -0.42 | (1.71) |
| COPD | -0.27 | (1.25) | 0.04 | (1.28) | -0.04 | (1.28) |
| Physicians | -1.57* | (0.74) | -1.97** | (0.74) | -2.05* | (0.89) |
| Hospital beds | -0.67 | (0.55) | -0.37 | (0.54) | -0.42 | (0.52) |
| Pharmacies | -0.11 | (0.37) | -0.04 | (0.38) | -0.23 | (0.45) |
| Infant mortality | 0.53 | (0.66) | 0.49 | (0.65) | 0.47 | (0.64) |
| Life expectancy | 7.22** | (2.42) | 6.12* | (2.49) | 6.85* | (3.38) |
| Need of care | 0.00 | (0.03) | 0.00 | (0.03) | 0.00 | (0.03) |
| Region |  |  |  |  |  |  |
| Pop. density | 0.01 | (0.00) | 0.01† | (0.00) | 0.01† | (0.00) |
| Car density | 0.02 | (0.03) | 0.02 | (0.03) | 0.02 | (0.03) |
| Commuter balance | 0.09 | (0.09) | 0.06 | (0.09) | 0.08 | (0.08) |
| Share of foreigners | -0.13 | (0.61) | -0.25 | (0.61) | -0.07 | (0.64) |
| Multi-fam. houses | 0.21 | (0.22) | 0.26 | (0.22) | 0.27 | (0.22) |
| Region type | 1.55 | (2.33) | 0.43 | (2.44) | 0.82 | (2.50) |
| Pupils | -2.09† | (1.16) | -2.21* | (1.11) | -1.91† | (1.12) |
| Childcare | -0.48† | (0.28) | -0.41 | (0.29) | -0.52† | (0.29) |
| Constant | -923.01** | (252.08) | -848.56** | (254.05) | -942.06 | (.) |
| lambda | -0.31† | (0.18) |  |  | -0.47* | (0.21) |
| rho | 0.44** | (0.02) | 0.79* | (0.35) |  |  |
| sigma2 | 446.89** | (31.61) | 437.83** | (30.98) | 430.65** | (32.05) |

In the area of health indicators it is noticeable that the prevalence of widespread diseases such as COPD and heart failure has no significant influence on the number of cases on an ecological level. The fact that this ecological result cannot, of course, be transferred to the individual level in individual cases is only repeated here, cf. [32] for first results of a significant positive significant association of a severe course of COVID-19 with COPD at individual level. However, a high regional physician density seems to have a significantly reducing effect on the number of cases. This may indicate, for example, an early implementation of quarantine and hygiene measures favoured by the presence of physicians. However, this correlation apparently does not apply to hospital beds and pharmacies.

With regard to regional characteristics, as expected, population density appears to have a significant (1% level) impact on the incidence of infection, which means empirical confirmation of common epidemiological models of virus spread and also corresponds to public perception. Here, no further significant correlation seems to emerge when looking at the housing situation in multi-family houses. Nevertheless, the result is important, as it seems to exclude a possible channel, at least in this empirical snapshot, at the ecological level. The same applies to car density (also as a proxy for air quality), commuter balance and the proportion of foreigners. The central result in this area for our research question No. 1, however, are the significantly negative coefficients for the number of schoolchildren as well as the childcare rate for infants (the latter is significant only in the SAR model). In this ecological study (in contrast to the predominant public discussion), the density of pupils seems to have not only no positive, but even a negative influence on the incidence of infection. Of course, this does not mean that schools can be excluded as hotspots for infection chains in individual cases. However, it does call for further research in this area.

### 3.2 Death count

The death rates in relation to COVID-19 were also investigated in analogy to the case numbers, using the spatial models discussed in Section 2.2. The results are summarized in Table 3. In comparison with the results for the case numbers in Table 2, it is noticeable that the signs and significances for the estimated coefficients are virtually identical. To explain this, it should be taken into account that the death rates usually represent a percentage (mortality) of the case numbers with a certain time lag. For Germany, this mortality is currently (as of June 15, 2020) approx. 4.7%. However, a separate consideration of the death rates as outcome variables is nevertheless justified, since mortality (especially at the beginning of an epidemic) is by no means a time-constant variable and therefore the structural similarity of the socio-economic influencing variables compared to the case numbers is not known a priori.

### 3.3 Spillover effects

Regarding our question No. 3 stated in Section 1, the above LR tests in Section 3.1 already support the thesis of a spatial dependency in the data. However, the question of actual regional spillover effects remains unanswered. In [9] it is proposed to test the presence of these spatial spillover effects via the so-called indirect effects. The latter reflect how a change in the *k*-th independent variable of region *j* affects the number of cases (or deaths) in region *i*, as the result of an infinite sequence of feedback effects. Since these effects naturally differ regionally, summary measures were proposed as quasi-average spillover effects. For technical details see for example [15].

In order to keep the presentation clear, in Table 4 we give only the summary measures of the significant indirect effects and the corresponding total effects. These allow us to interpret which socio-economic variables have different effects on neighbouring regions (spillovers) than they have in total (total effects, i.e. including direct effects within the same region). The SEM model has no spillover effects and is therefore omitted in Table 4.

**Table 4:**
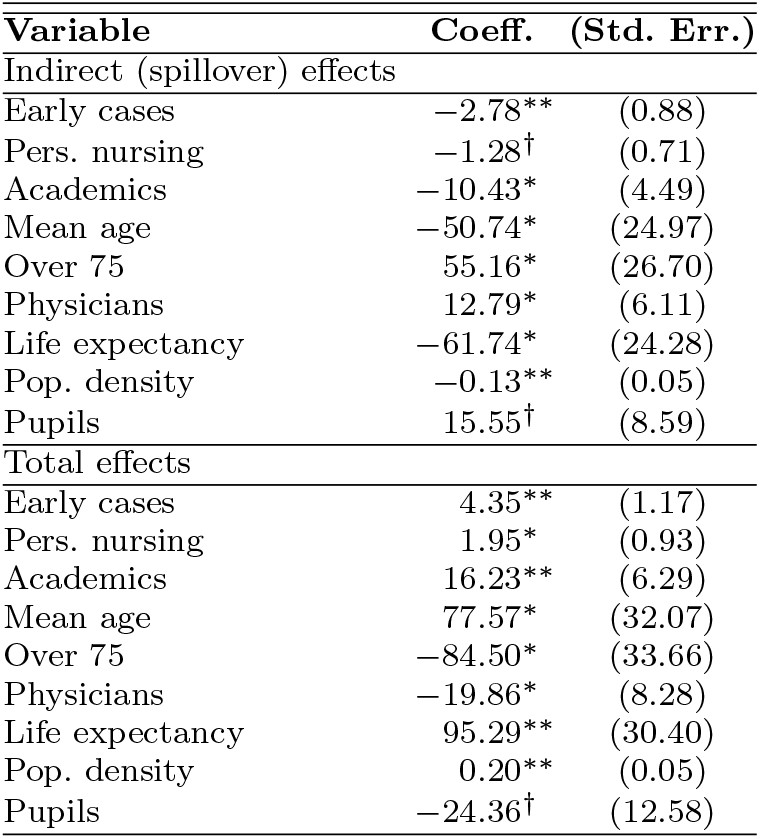
COVID-19 case numbers: Direct and indirect effects for the SAR model. The significance level symbols are as before.

| Variable | Coeff. | (Std. Err.) |
| --- | --- | --- |
| Indirect (spillover) effects |  |  |
| Early cases | -2.78** | (0.88) |
| Pers. nursing | -1.28<sup>†</sup> | (0.71) |
| Academics | -10.43* | (4.49) |
| Mean age | -50.74* | (24.97) |
| Over 75 | 55.16* | (26.70) |
| Physicians | 12.79* | (6.11) |
| Life expectancy | -61.74* | (24.28) |
| Pop. density | -0.13** | (0.05) |
| Pupils | 15.55<sup>†</sup> | (8.59) |
| Total effects |  |  |
| Early cases | 4.35** | (1.17) |
| Pers. nursing | 1.95* | (0.93) |
| Academics | 16.23** | (6.29) |
| Mean age | 77.57* | (32.07) |
| Over 75 | -84.50* | (33.66) |
| Physicians | -19.86* | (8.28) |
| Life expectancy | 95.29** | (30.40) |
| Pop. density | 0.20** | (0.05) |
| Pupils | -24.36<sup>†</sup> | (12.58) |

For an interpretation of the results in Table 4 only selected variables should be highlighted, because not in all cases the discussion of a potential spatial mechanism of action seems to be reasonable at the current state of research. For example, the significantly negative spillover effects of the early cases could indicate that early measures of lockdown in the affected regions as well as increased attention and thus caution towards COVID-19 have successfully contributed to a non-spillover of the cases to neighbouring regions. In the case of the also significantly negative spillover effects of the mean age, it could be argued that reduced travel to neighbouring regions and thus a containment effect on the number of cases there could be achieved. An interpretation of the significantly positive spillover effects for the proportion of over 75-year-olds, however, seems difficult. Further, it is interesting to note that the number of beds in tourist establishments does not seem to lead to significant spillover effects in neighbouring regions (not included in the table). Here, for example, the early restrictions on tourist travel in the context of the imposed contact restrictions could play a role. Also not significant, however, are car density and commuters, who were less affected by the contact restrictions. On the other hand, the density of doctors seems to lead to significant positive spillover effects. Here, the importation of infections through cross-county doctor visits could be discussed as an argument. The same (i.e. the import of infections) could possibly be considered as an explanation for the significantly positive spillover effects of pupils. However, the last two arguments should be considered with caution and as an indication of the need for further research. For death rates, we find no significant indirect effects on any of the socio-economic variables considered.

## 4 Discussion

This ecological study deals with the influence of socio-economic variables on the COVID-19 infection process. Data at the district level and spatial econometric models are used, e.g. to capture indirect spillover effects between the individual districts. In particular, it is a question of examining which measures outside the clinical area or the health care sector have an influence on the occurrence of infection. We had asked three questions at the beginning and found interesting hints in Section 3. For example, the density of pupils and the care rate of small children (kindergartens) seems to have a rather dampening (i.e. negative) effect on the infection rate. The same applies to the number of deaths. However, positive spillover effects of pupil density on neighbouring regions could be observed, so that this cross-circle export or import of infections should be analysed more closely in future research. Also on our second question, namely the role of care for the elderly and the employees in this area, first significant indications could be found. The results show that the sole quota of persons in need of care has no influence on infections and deaths, whereas the influence of the employment density in this sector on both outcomes turns out to be significantly increasing. These results are all the more interesting because they also support the anecdotal evidence from the media about nursing homes as hotspots of COVID-19 dissemination and even differentiate between the different outcomes for residents and employees. However, it must be mentioned as a limitation that our data do not distinguish between day care at home and nursing homes. We were also able to find clear evidence for our third question of spillover effects between neighbouring districts, which, to our knowledge, was investigated empirically for COVID-19 in Germany for the first time here. On the one hand, the suitability of spatial models compared to simple OLS approaches is statistically supported. On the other hand, indications of negative effects (i.e. a containment effect on neighbouring regions) could be found especially for the number of early cases of infection or the average age, whereas spillover effects for doctor and pupil density turn out to be positive. Since this is a quasi export of infections across national borders, these questions should be analysed in future studies in greater detail and, if possible, on an even smaller regional scale, in order to expand the knowledge about the ecological pathways of COVID-19. In addition to the three above-mentioned questions, the counterintuitive positive influence of the economic status (e.g. household income or proportion of acedemics) of the districts on the occurrence of infections, as known from other studies, could be empirically confirmed.

The practical relevance of the results lies, among other things, in the derivation of new or adaptation of existing political measures for the containment of COVID-19, which by their very nature also do not function at the individual level, but at the population level. When interpreting these ecological results, different mechanisms of action, namely the numerous interdependent infection channels at population level, must be taken into account than in clinical or biological studies at individual level. To give one example: Certainly, a high social status, which can be sufficiently measured e.g. by income, at least statistically protects against infection or even death from COVID-19 at the individual level. Well-known mechanisms of action from the literature can include higher education associated with income, better access to medical care or an information advantage. At the population level, however, this effect can be reversed (especially in the initial phase of the epidemic). A high level of economic activity is often based on networking (including physical networking), travel and social contacts such as working with colleagues in teams or open-plan offices. In other words, both results can be empirically substantiated and are therefore not contradictory.

It is precisely here that the identification of population-based risk groups helps policymakers to develop targeted and effective strategies that go beyond the purely clinical sphere of influence. Concrete impulses could arise from this analysis, among other things with regard to a possible shift in focus of contact restrictions. It would be conceivable to discuss, for example, the tourism sector as well as schools and infant care, none of which could be identified in our study as drivers of infection or death rates. However, it should be taken into account that this study is a snapshot and, in particular, no causalities can be deduced about the effect of existing contact restrictions. In other words, it cannot be excluded in the context of this study that early school closures in particular contributed to the dampening effect of pupil density on infection rates. Future research could build on this and include the exact regional measures for contact restriction in their chronological sequence in an extended analysis.

## Data Availability

All data used are publicly available.

## Competing interests

The author declares that he has no competing interests nor financial relationships with any organizations that might have an interest in the submitted work. This research did not receive any specific grant from funding agencies in the public, commercial, or not-for-profit sectors.

## Footnotes

1 Called up on 11.06.2020 at https://public.opendatasoft.com/explore/?sort=modified

2 For details, see e.g. https://www.bundesregierung.de/breg-de/themen/coronavirus/corona-bundeslaender-1745198

## Notes

### Competing Interest Statement

The authors have declared no competing interest.

### Author Declarations

No person-specific data was used in this study.

